# Deep neural networks detect regional wall motion abnormalities and preclinical cardiovascular disease from 12-lead ECGs

**DOI:** 10.1101/2024.05.31.24308304

**Authors:** Tanner Carbonati, Parastou Eslami, Jonathan W. Waks, Laurent Fiorina, Ashish Chaudhari, Christine Henry, Alistair E.W. Johnson, Tom Pollard, Brian Gow, Roger G. Mark, Steven Horng, Nathaniel R. Greenbaum

## Abstract

**Background:** Identifying regional wall motion abnormalities (RWMAs) is critical for diagnosing and risk stratifying patients with cardiovascular disease, particularly ischemic heart disease. We hypothesized that a deep neural network could accurately identify patients with regional wall motion abnormalities from a readily available standard 12-lead electrocardiogram (ECG).

**Methods:** This observational, retrospective study included patients who were treated at Beth Israel Deaconess Medical Center and had an ECG and echocardiogram performed within 14 days of each other between 2008 and 2019. We trained a convolutional neural network to detect the presence of RWMAs, qualitative global right ventricular (RV) hypokinesis, and varying degrees of left ventricular dysfunction (left ventricular ejection fraction [LVEF] ≤50%, LVEF ≤40%, and LVEF ≤35%) identified by echocardiography, using ECG data alone. Patients were randomly split into development (80%) and test sets (20%). Model performance was assessed using area under the receiver operating characteristic curve (AUC). Cox proportional hazard models adjusted for age and sex were performed to estimate the risk of future acute coronary events.

**Results:** The development set consisted of 19,837 patients (mean age 66.7±16.4; 46.7% female) and the test set comprised of 4,953 patients (mean age 67.5±15.8 years; 46.5% female). On the test dataset, the model accurately identified the presence of RWMA, RV hypokinesis, LVEF ≤50%, LVEF ≤40%, and LVEF ≤35% with AUCs of 0.87 (95% CI 0.858-0.882), 0.888 (95% CI 0.878-0.899), 0.923 (95% CI 0.914-0.933), 0.93 (95% CI 0.921-0.939), and 0.876 (95% CI 0.858-0.896), respectively. Among patients with normal biventricular function at the time of the index ECG, those classified as having RMWA by the model were 3 times the risk (age- and sex-adjusted hazard ratio, 2.8; 95% CI 1.9-3.9) for future acute coronary events compared to those classified as negative.

**Conclusions:** We demonstrate that a deep neural network can help identify regional wall motion abnormalities and reduced LV function from a 12-lead ECG and could potentially be used as a screening tool for triaging patients who need either initial or repeat echocardiographic imaging.

## Introduction

Ischemic heart disease (IHD) remains the leading cause of death and disability worldwide^1,2^. IHD causes a wide spectrum of symptoms, and some patients with undiagnosed coronary artery disease may present with atypical or without obvious cardiac symptoms, which may delay diagnosis and intervention. Once identified, interventions such as medications and revascularization can be initiated to improve patient outcomes^3^. Echocardiography is an important non-invasive diagnostic tool for identifying patients with IHD through its ability to identify global ventricular dysfunction and regional wall motion abnormalities (RWMAs) that correlate with prior myocardial infarction or areas with hibernating myocardium due to inadequate blood flow. However, echocardiography is expensive, time consuming, and requires expert interpretation. Assessment of RWMA can help in the diagnosis of acute and chronic myocardial infarction^4^, as well as differentiating between ischemic and non-ischemic cardiomyopathy.

Echocardiography is the most common diagnostic modality to determine RWMAs, however, it is unclear when initial or repeated imaging should be performed, as ischemic disease progresses at different rates. Furthermore, access to echocardiography is limited in many clinical settings due to lack of resources or expertise. The electrocardiogram (ECG), however, is a widely available and inexpensive diagnostic tool that is commonly used to evaluate patients with suspected acute coronary syndrome. Recent studies have demonstrated the ability of deep learning to detect left ventricular dysfunction, myocardial infarction, and many other cardiovascular diseases from the ECG^5–8^.

In this observational, retrospective study we developed and validated a novel deep learning model to identify the presence of RWMA, global right ventricular (RV) hypokinesis and reduced left ventricular ejection fraction (LVEF) based on analysis of ECG alone. We leveraged two open-access databases of matched patient ECG and echocardiography reports collected from the intensive care unit and emergency room^9,10^. In patients where our model detected RWMA, we explored the model’s ability to localize the abnormal regions of the myocardium. We also evaluated the model’s ability to identify patients with normal biventricular function at the time of initial screening, but at increased risk of future wall motion abnormalities, reduced LVEF, and acute coronary events.

## Methods

### Study design and participants

Digitally stored 12-lead ECGs from patients aged 18 years or older that were captured within 14 days of the patient having two-dimensional echocardiographic imaging performed at Beth Israel Deaconess Medical Center (BIDMC) between Jan 1, 2008, and Dec 31, 2019, were identified. In the case of a patient having more than one echocardiogram performed during the study period, all were included for model development and only the first available exam was used for testing. If a patient had more than one ECG within 14 days of an echocardiogram during the study period, all were included for model development and the ECG performed closest to the time of echocardiography was used for testing. Patients without assessment of LV function reported in the echocardiography notes were excluded (Figure 1).

**Figure 1.**
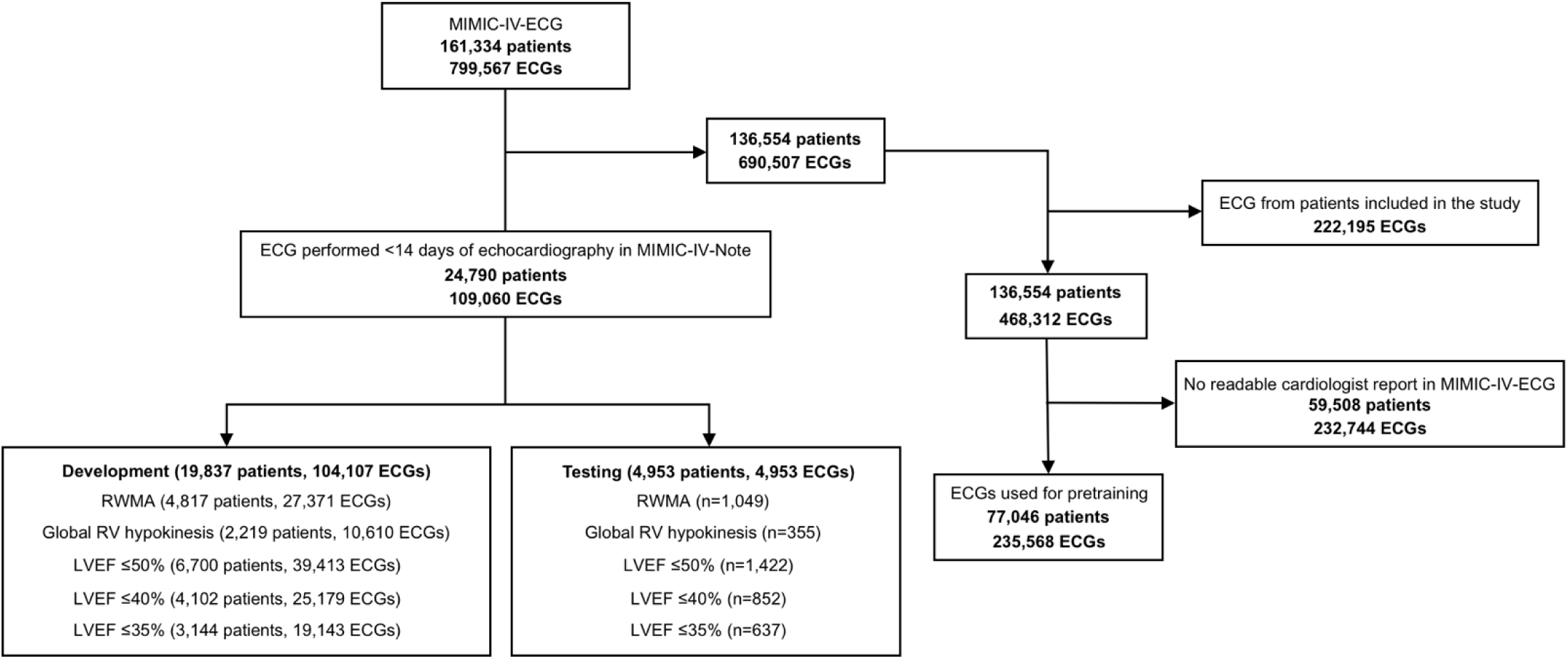
Study flow chart.

Data used for the study was collected from three public databases, Medical Information Mart for Intensive Care (MIMIC)-IV-ECG, MIMIC-IV-Note, and MIMIC-IV-ECHO^9–11^. Clinical data linked to patients included in the study was matched using the publicly available database, MIMIC-IV^12^.

### Ground truth

ECGs were labeled using echocardiography reports collected from the MIMIC-IV-Note dataset. RWMAs and their severity (hypokinesis, akinesis, or dyskinesis) were assigned to a location on the standard AHA 17-segment model^13^. All echocardiograms were read by a BIDMC cardiologist who was board certified in echocardiography. In the present study, we divided the left ventricle into seven regions in accordance with the coronary perfusion territories as: anterior, anterolateral, anteroseptal, apical, inferior, inferolateral, and inferoseptal^14^ (Supplementary Figure 1). We annotated the seven regions accordingly and included a composite RWMA endpoint for each patient defined as having ≥1 abnormal wall segment.

To assess global LV function, we included three clinically relevant LVEF endpoints as: LVEF ≤50%, LVEF ≤40%, and LVEF ≤35%^15^. Global RV hypokinesis was defined as qualitative global RV free wall hypokinesis.

### Data partition

Patients were randomly split into development (80%) and test sets (20%). The development set was used to train and validate the model using fivefold cross-validation. The first four folds were used to assess model robustness and the fifth fold was used to select the highest performing model and optimal thresholds. The test set consisted of patients not included in the development set and was used to report the model performance.

### Model development

We developed a multilabel convolutional neural network (CNN) to identify the presence of RWMAs, global RV hypokinesis, and reduced LVEF at various cut points from the 12-lead ECG. The model consisted of a feature-extraction component and classifier. For feature-extraction, we used a CNN encoder based on the resnet-101 architecture^16^ with lead-specific convolutions. The lead-specific convolutions allowed the network to extract temporal features from each lead in parallel. A final convolutional layer was used to aggregate the lead-specific features, which were passed to a global average pooling layer for classification. The classifier was composed of two fully connected layers with dropout before each layer, and a sigmoid activation applied to each output. The model takes as input a 10-second 12-channel ECG waveform with a sampling frequency of 500 Hz and outputs a probability score for the presence of RWMA with a score for each region (anterior, anterolateral, anteroseptal, apical, inferior, inferolateral, and inferoseptal), ≥1 akinetic or dyskinetic wall, LVEF ≤50%, LVEF ≤40%, and LVEF ≤35%, and global RV hypokinesis. Detailed processes for model development and hyperparameter tuning are provided in the appendix.

For comparison, a reference model was derived using patient demographics and ECG findings extracted from cardiologist reports available in MIMIC-IV-Note. We trained a random forest classifier using as input the patient age, sex, and presence of 41 different ECG findings (Supplementary Table 1).

Furthermore, we pretrained the deep learning model on patients from the MIMIC-IV-ECG dataset without an associated echocardiography report available in MIMIC-IV-Note (Figure 1). The model was pre-trained to detect the presence of 41 different ECG findings extracted from the ECG cardiologist overreads available in MIMIC-IV-Note. All findings were extracted from the reports using regular expressions. An overview of the ECG findings used for pre-training are provided in the appendix (Supplementary Table 1). No patients included in the study were used for pre-training (Figure 1).

### Model evaluation

The primary objective of the study was to evaluate the ability of a deep neural network to identify any RWMA, reduced LVEF, and global RV hypokinesis from a 12-lead ECG using the MIMIC-IV-ECG and MIMIC-IV-Note datasets. We additionally assessed performance of the model to localize RWMA classification to one or more of the seven regions of the left ventricular myocardium reflective of coronary perfusion territories. To measure potential bias from the model we performed subgroup analyses across patient age, sex, and self-reported race/ethnicity found in the MIMIC-IV dataset and cardiologist reported ECG findings. Diagnostic odds ratios (ORs) were computed for each subgroup.

Our secondary objective was to assess the prognostic value of the model to identify patients with normal biventricular function at the time of their index ECG-echocardiogram, but at increased risk of developing future RWMAs, reduced LVEF, RV hypokinesis, and a composite label for any of the endpoints during 5-year follow-up. Subsequent echocardiograms following the index ECG-echocardiogram were used for analysis of follow-up. We also evaluated the model’s ability to identify patients with normal biventricular function and no history of ischemic heart disease diagnosed prior to or during their admission of screening, but at increased risk of acute coronary events during 5-year follow-up. Acute coronary events were defined as myocardial ischemia or its acute complications, or a coronary revascularization procedure such as percutaneous angioplasty/stent placement or coronary artery bypass graft surgery. Clinical outcomes and acute coronary events were identified by the presence of International Classification of Diseases (ICD)-10 and ICD-9 codes available in MIMIC-IV^12^ (Supplementary Table 2). Cox proportional hazard models adjusted for age and sex were performed to estimate the risk of each endpoint.

### Statistical analysis

The performance of the model was evaluated using the area under the receiver operating characteristic curve (AUC), sensitivity, specificity, positive and negative predictive values. To estimate 95% confidence intervals (CIs), we used non-parametric bootstrapping with 1,000 samples. Sensitivity and specificity were calculated at binary decision thresholds. The optimal decision threshold for each class was calibrated using the receiver operating curve and Youden index analysis on the final cross-validation fold. Delong’s test was used to compare the AUC between the deep learning model and reference model on the test set. Continuous variables were compared using Student’s t-test and categorical variables were compared with chi-Squared test. We considered two-sided p values <0.05 statistically significant. Kaplan-Meier analysis was used to compare the incidence of RWMA, global RV hypokinesis, reduced LVEF, and acute coronary event for true negatives versus false positives during follow-up. All models and statistics were computed using Python (v.3.8.12). Deep learning models were trained using the PyTorch (v1.7.0). Data analysis was performed using numpy (1.19.5), pandas (1.2.0), scipy (1.6.0), and scikit-learn (0.24.0). For data visualization and scientific plotting matplotlib (3.2.2) and seaborn (0.12.2) were used. Kaplan-Meier curves were computed using R (v 4.3.1) with the survival and survminer packages.

## Results

A total of 109,060 ECGs associated with 39,144 echocardiograms from 24,790 patients were included in the study. The development set consisted of 19,837 patients (mean age 66.7±16.4 years; 46.7% female) and the test set consisted of 4,953 patients (mean age 67.5±15.8 years; 46.5% female). Median time between ECG and echocardiography of the training set and test sets were 1.8 days (IQR 0.7-4.7 days) and 0.77 days (IQR 0.2-1.8 days). A RWMA was observed in 3856 (19.4%) and 1049 (21.2%) of patients in development and test sets and global RV hypokinesis in 1360 (6.9%) and 355 (7.2%), respectively (Table 1). Sample sizes for segment level wall motion abnormalities are provided in Supplementary Table 3 and baseline ECG findings in Supplementary Table 1.

**Table 1.** Patient baseline characteristics at the time of the index ECG-echocardiogram. Values are n (%) or mean SD. Student t-test and chi-squared test were used (BMI=body mass index, LVEF=left ventricular ejection fraction, RWMA=regional wall motion abnormality, RV=right ventricular)

|  | **Train** | **Test** | **P value** |
| --- | --- | --- | --- |
| **Patients** | 19837 | 4953 |  |
| **Age** | 67.5 ± 15.6 | 66.5 ± 16.6 | <0.001 |
| **Female** | 9067 (45.7) | 2345 (47.3) | 0.04 |
| **White** | 13394 (67.5) | 3260 (65.8) | 0.024 |
| **Black/African American** | 2316 (11.7) | 655 (13.2) | 0.003 |
| **Hispanic** | 747 (3.8) | 191 (3.9) | 0.797 |
| **Asian** | 492 (2.5) | 161 (3.3) | 0.003 |
| **Other** | 26 (0.1) | 7 (0.1) | 0.968 |
| **Unknown or choose not to disclose** | 2861 (14.4) | 678 (13.7) | 0.194 |
| **BMI** | 33.5 ± 134.7 | 28.9 ± 55.3 | 0.062 |
| **LVEF** | 54.0 ± 21.0 | 53.6 ± 12.9 | 0.361 |
| **Heart Failure** | 2098 (10.6) | 395 (8.0) | <0.001 |
| **Myocardial Infarction** | 1223 (6.2) | 231 (4.7) | <0.001 |
| **Hypertension** | 6637 (33.5) | 1464 (29.6) | <0.001 |
| **Coronary artery disease** | 2886 (14.5) | 579 (11.7) | <0.001 |
| **Diabetes** | 3040 (15.3) | 621 (12.5) | <0.001 |
| **RWMA** | 3856 (19.4) | 1049 (21.2) | 0.006 |
| **Global RV hypokinesis** | 1360 (6.9) | 355 (7.2) | 0.744 |
| **LVEF ≤50%** | 5511 (27.8) | 1422 (28.7) | 0.259 |
| **LVEF ≤40%** | 3234 (16.3) | 852 (17.2) | 0.164 |
| **LVEF ≤35%** | 2424 (12.2) | 637 (12.9) | 0.268 |

On the test dataset, the deep learning model accurately identified the presence of any RWMA with an AUC of 0.87 (95% CI 0.858-0.882), a sensitivity of 74.6 (95% CI 72.1-77.4), and a specificity of 82.3 (95% CI 81.3-83.5) (Table 2). For comparison, the reference model yielded an AUC, sensitivity, and specificity of 0.70 (95% CI 0.682-0.724), 78.4 (95% CI 81.1-75.7), and 48.4 (95% CI 45-51.8), respectively (Supplementary Table 4). The deep learning model significantly outperformed the reference model to detect RWMA (P<0.001). The deep learning model identified abnormal wall motion in individual wall segments with an AUC ranging from 0.853 to 0.913, sensitivity from 69.3 to 83.4%, and specificity from 76.7 to 83.8% (Table 2). The sensitivity of the model to identify RWMA was correlated with the number of abnormal wall segments (Supplementary Figure 2).

**Table 2.** Model performance of the deep learning model on the test set. (RWMA=regional wall motion abnormality, LVEF=left ventricular ejection fraction, RV=right ventricular, AUC=area under the receiver-operator curve, PPV=positive predictive value, NPV=negative predictive value, 95% CIs computed using bootstrapping with 1,000 samples)

|  | AUC | Sensitivity | Specificity | PPV | NPV |
| --- | --- | --- | --- | --- | --- |
| **RWMA** | 0.87 (0.858-0.882) | 74.6 (72.1-77.4) | 82.3 (81.3-83.5) | 53.2 (50.7-55.9) | 92.4 (91.4-93.3) |
| **LVEF $\leq 50\%$** | 0.888 (0.878-0.899) | 74.7 (72.4-76.8) | 86.8 (85.7-87.9) | 69.7 (67.4-71.9) | 89.4 (88.4-90.4) |
| **LVEF $\leq 40\%$** | 0.923 (0.914-0.933) | 86.7 (84.5-89.0) | 83.5 (82.3-84.6) | 52.3 (49.7-54.9) | 96.8 (96.2-97.3) |
| **LVEF $\leq 35\%$** | 0.93 (0.921-0.939) | 83.0 (80.0-86.0) | 86.6 (85.6-87.6) | 48.0 (45.0-50.9) | 97.2 (96.7-97.7) |
| **Global RV hypokinesis** | 0.876 (0.858-0.896) | 74.9 (70.2-79.1) | 84.4 (83.3-85.4) | 28.2 (25.6-31.1) | 97.6 (97.1-98.1) |
| **Anterior wall** | 0.913 (0.9-0.926) | 81.6 (77.5-85.6) | 83.8 (82.7-84.9) | 28.6 (26.0-31.3) | 98.3 (97.9-98.7) |
| **Anterolateral wall** | 0.89 (0.874-0.908) | 81.1 (76.0-86.5) | 79.8 (78.8-81.0) | 15.8 (13.8-18.1) | 98.9 (98.6-99.2) |
| **Anteroseptal wall** | 0.896 (0.881-0.911) | 82.2 (78.3-85.7) | 82.1 (81.1-83.3) | 29.4 (26.8-32.0) | 98.1 (97.6-98.5) |
| **Apex** | 0.894 (0.882-0.906) | 83.4 (80.2-86.1) | 79.2 (78.1-80.5) | 37.4 (35.0-39.9) | 97.0 (96.4-97.5) |
| **Inferior wall** | 0.854 (0.838-0.869) | 79.6 (76.4-82.5) | 76.7 (75.5-77.9) | 36.6 (34.1-39.1) | 95.7 (94.9-96.4) |
| **Inferolateral wall** | 0.853 (0.836-0.869) | 69.3 (65.5-73.2) | 83.4 (82.5-84.5) | 36.2 (33.5-39.3) | 95.2 (94.5-96.0) |
| **Inferoseptal wall** | 0.878 (0.862-0.894) | 82.6 (78.4-86.6) | 77.5 (76.5-78.8) | 22.4 (20.3-24.8) | 98.3 (97.8-98.7) |
| **$\geq 1$ hypokinetic wall** | 0.833 (0.819-0.849) | 72.2 (69.4-75.3) | 78.8 (77.6-80.1) | 41.4 (39.0-44.2) | 93.2 (92.3-94.1) |
| **$\geq 1$ akinetic wall** | 0.905 (0.892-0.917) | 85.2 (82.1-88.0) | 80.4 (79.3-81.6) | 34.7 (32.4-37.4) | 97.8 (97.3-98.3) |

The deep learning model accurately detected global RV hypokinesis with an AUC of 0.876 (0.858-0.896) (Table 2). The deep learning model also accurately identified LVEF ≤50%, LVEF ≤40%, and LVEF ≤35% with AUCs of 0.888 (95% CI 0.878-0.899), 0.923 (95% CI 0.914-0.933), and 0.93 (95% CI 0.921-0.939), respectively. For the composite of any RMWA or LVEF ≤50% the model yielded a sensitivity of 82.7% with a positive predictive value of 64.6%.

We evaluated model performance across subgroups of patient age, sex, and race/ethnicity (Table 3). The model showed consistent performance for both sexes. Performance was robust across all age groups for the detection of reduced LVEF and RV hypokinesis, however, model performance to identify RMWAs was comparatively lower in older patients compared to younger patients. We observed no significant difference in performance across patient race/ethnicity for each abnormality. Notably, we did not have sufficient data to test patients of Hispanic or Asian race/ethnicity independently, however, when grouped (Hispanic n=191, age 66.6 ± 16.6, 43.5% female; Asian n=161, age 66.6 ± 16.6, 43.5% female; American Indian n=7, age 61.6 ± 19.7, 57.1% female) during analysis we observed favorable performance of the model. Additional subgroup analysis was performed across ECG findings (Supplementary Figure 3) with similar model performance regardless of ECG findings apart from paced rhythms or left bundle branch block where model performance was slightly reduced.

Among patients with normal biventricular function at the time of the index ECG, those classified as having a RWMA (false positive) by the model were 3 times the risk (age- and sex-adjusted hazard ratio, 3.2; 95% CI 2.2-4.6) of presenting with a future RWMA compared to those initially classified as negative (true negative) (Figure 2A). Similarly, the hazard ratios comparing false positives and true negatives for global RV hypokinesis and LVEF ≤40% were 6.2 (95% CI 3.6-10.8) and 4.1 (95% CI 2.7-6.4), respectively (Figure 2B-C). Patients classified as positive for at least one abnormality by the model were 3 times the risk (3.0 HR; 95% CI 2.3-4.0) of presenting a future composite event (RWMA, global RV hypokinesis, or LVEF ≤50%). Among patients with normal biventricular function and no history of ischemic heart disease, patients classified as positive for RWMA had a 2.8-fold increased hazard for acute coronary events (95% CI 1.9-3.9) (Figure 3).

**Figure 2.**
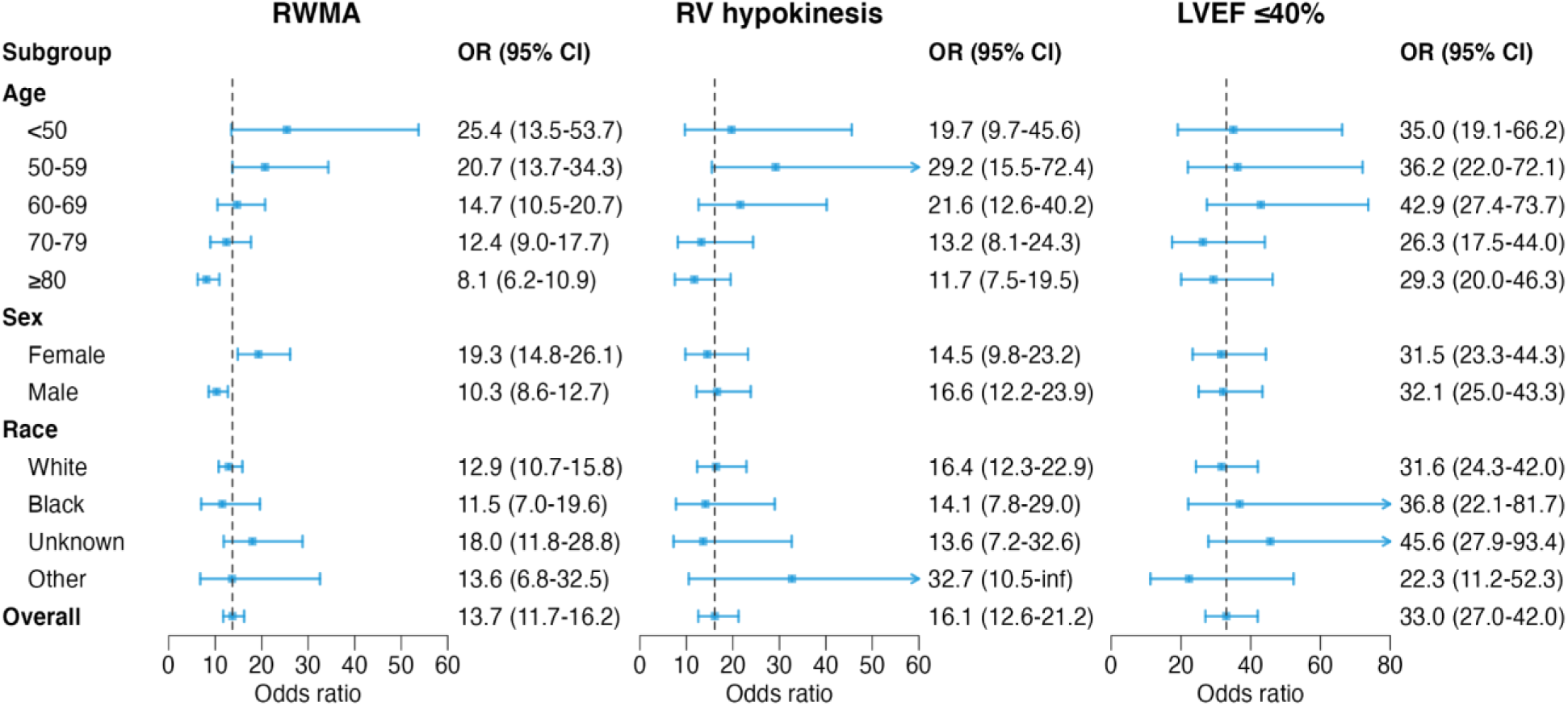
Subgroup analysis of the deep learning model by patient age, sex, and race/ethnicity using diagnostic odds ratio (OR) with 95% confidence intervals (CIs). The vertical dashed lines represent the OR of the model across all patients in the test set.

**Figure 3.**
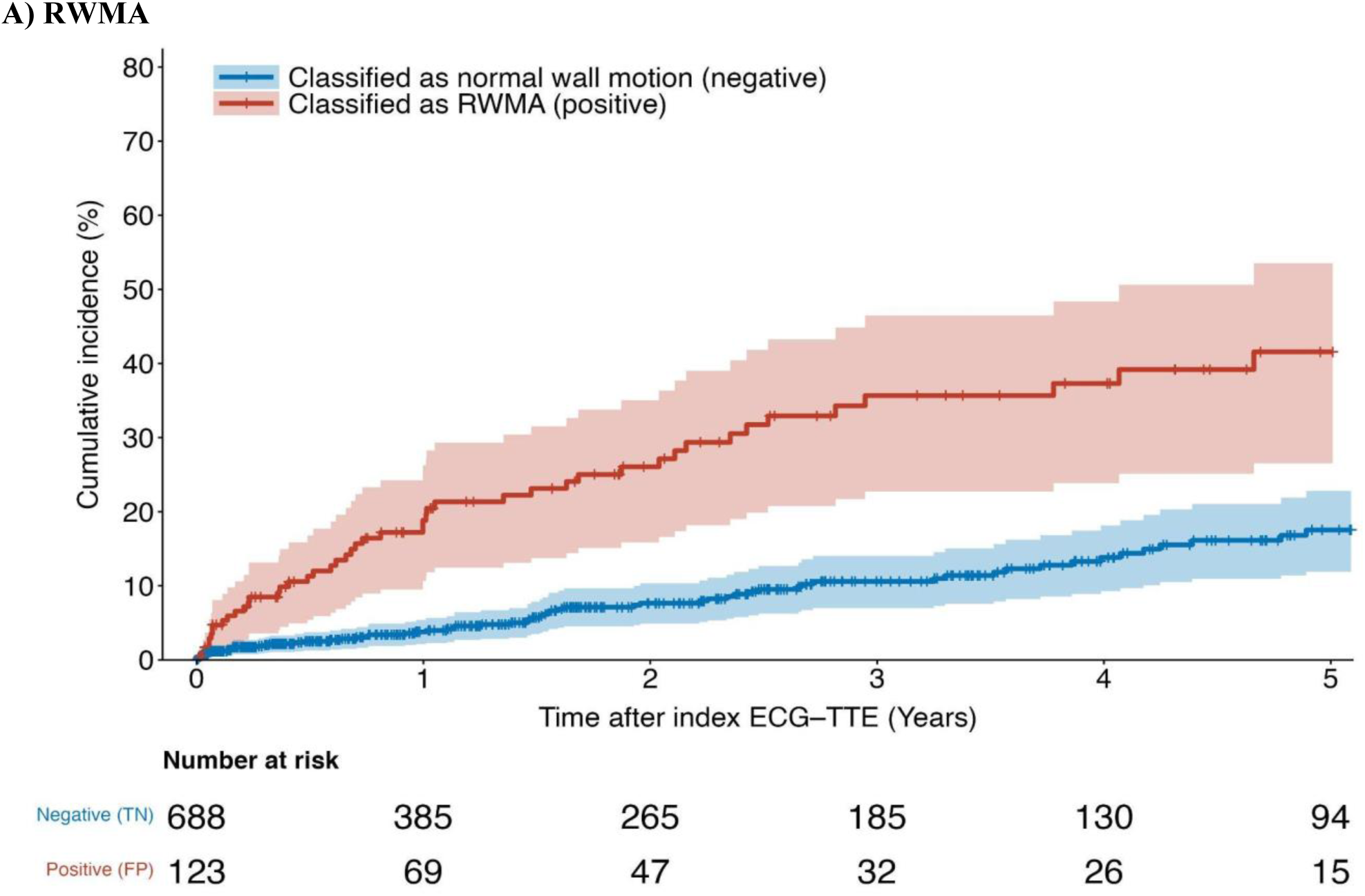
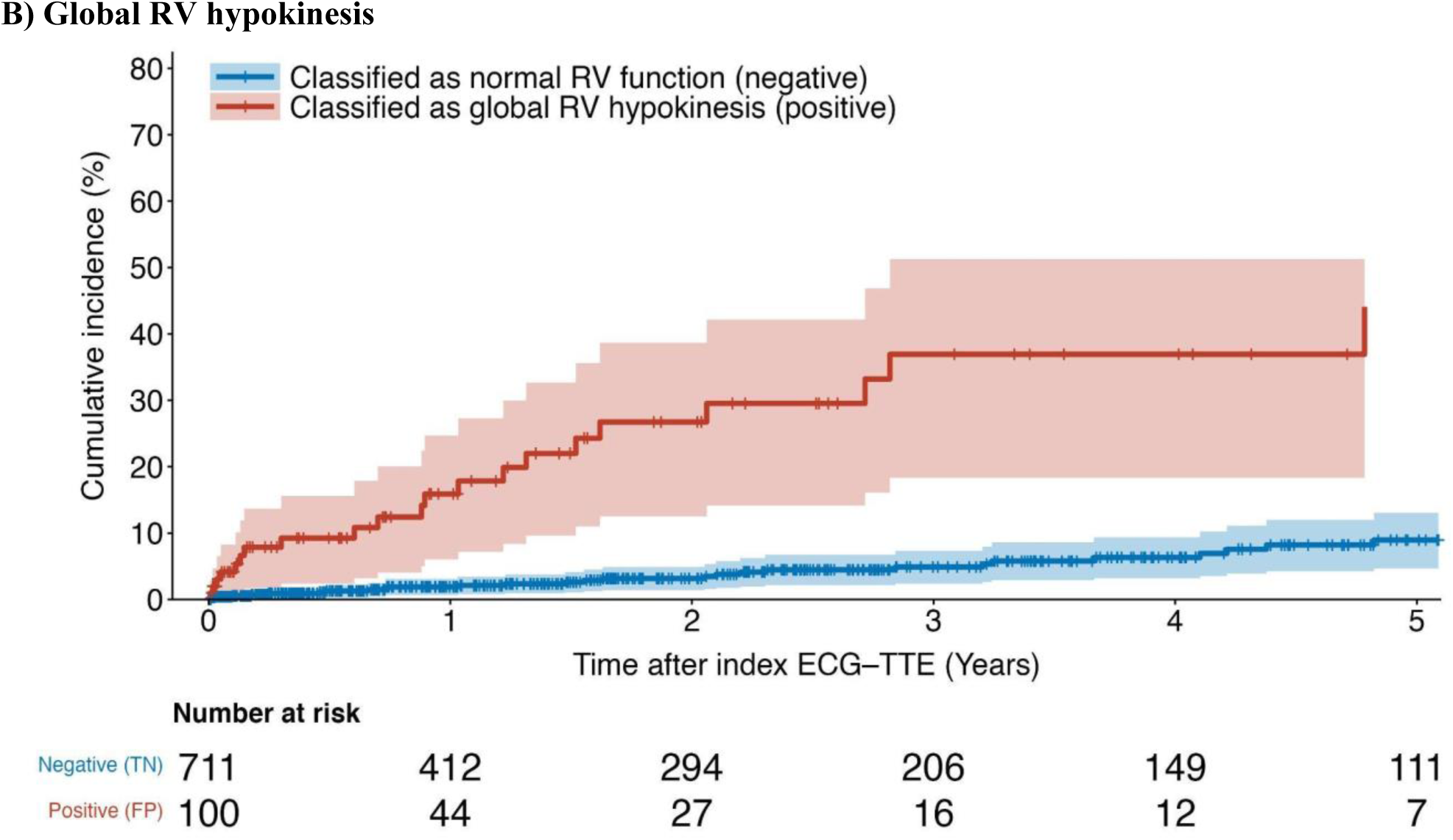
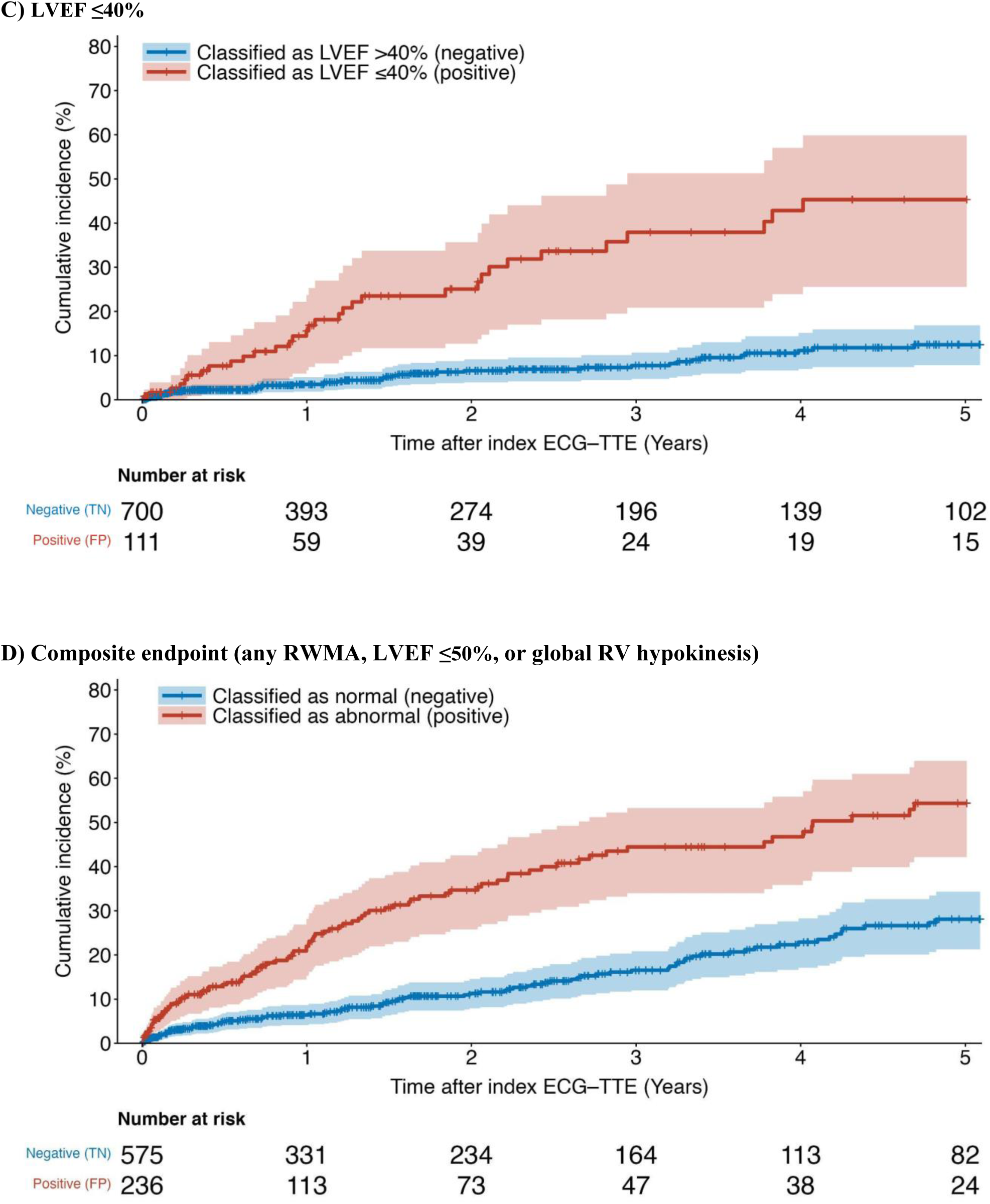
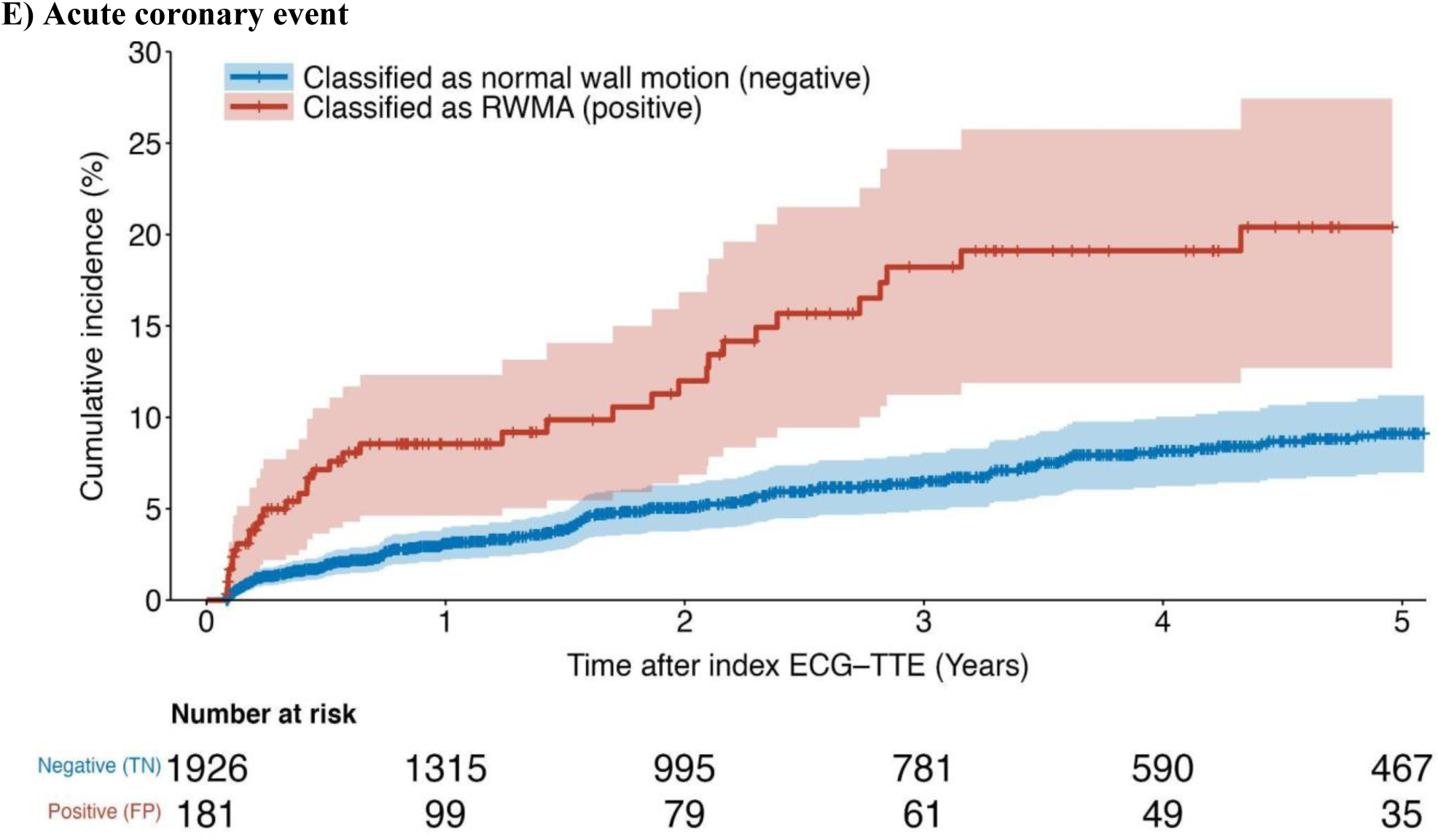
Cumulative incidence of cardiovascular outcomes in patients with initially normal biventricular function at the time of the index ECG-echo stratified by model classification during a 5-year follow-up period. **(A)** Cumulative incidence curves for RWMA stratified by classification of RWMA **(B)** Cumulative incidence curves for global RV hypokinesis stratified by classification of global RV hypokinesis. **(C)** Cumulative incidence curves for LVEF ≤40% stratified by classification of LVEF ≤40% **(D)** Cumulative incidence curves for a composite endpoint (any RWMA, LVEF ≤50%, or global RV hypokinesis) stratified by any abnormal classification (**E**) Cumulative incidence curves for acute coronary events in patients with normal biventricular function and no history of ischemic heart disease prior to or during the index admission stratified by classification of RWMA.

## Discussion

In summary, we developed and validated a novel deep learning model to identify and localize regional motion abnormalities from a 12-lead ECG alone. The added value of this study is twofold: First, we show that deep neural networks can accurately detect abnormal wall motion in individual wall segments (AUC: 0.85 to 0.91), global RV hypokinesis (AUC: 0.88), LVEF ≤50% (AUC: 0.89) LVEF ≤40% (AUC: 0.92), and LVEF ≤35% (AUC: 0.93). Second, we showed that our model may be able to detect subtle patterns in the ECG able to identify patients with normal biventricular function at the time of initial screening, but at increased risk of future ischemic heart disease (Figure 3). Early identification of high-risk patients may enable healthcare providers to intervene proactively, preventing the occurrence of detrimental events such as suffering a heart attack.

This study is the first to accurately identify and localize regional wall motion abnormalities from the 12-lead ECG. While recent studies have demonstrated the potential of deep learning-enabled ECGs to detect severely reduced LVEF, a large proportion of these studies were developed and validated using closed or proprietary datasets^5–8^. Attia et al demonstrated that deep neural networks can detect LVEF ≤35% from the 12-lead ECG with an AUC of 0.94. Similarly, Vaid et al detected LVEF at cutoffs of 50%, 40%, and 35% with similar AUCs of 0.89, 0.94, and 0.95, respectively.

Our study adds to this previous work for the detection of reduced LVEF by introducing a model that explicitly identifies the presence of RWMAs, which may help differentiate if the reduced LVEF may be from a focal abnormality compared to a global disease, and which has important implications for further diagnosis and intervention. We are unable to compare the performance of our model to prior studies because the models and data are unavailable.

Our choice of dividing the left ventricle into seven myocardial regions reflective of the coronary perfusion territories may provide value in identifying patients with single-vessel versus multivessel coronary artery disease. These seven regions are also in accordance with ASE segmentation guidelines^14^ and may complement RWMA assessment during echocardiography, especially when views are suboptimal. Additionally, our model could be used to detect which patients require repeat electrocardiography. For patients with suspected acute coronary syndrome, this model may help monitor both the onset of new RWMAs or changes in LVEF that require further evaluation or intervention.

In addition to effectively identifying patients with RWMA or reduced LVEF, our secondary objective demonstrates that our deep learning model may also identify patients with seemingly normal biventricular function who are at increased risk of future RWMA. Notably, these patients predicted as positive by model (false positive) had a threefold increased risk of developing a new RWMA over the next 5 years. We repeated this analysis for global RV hypokinesis and LVEF ≤40%, which revealed a sixfold and fourfold increased risk for each, respectively. In patients with normal biventricular function and no history of ischemic heart disease, the false positives of the model had a near threefold increased risk of acute coronary events. These findings suggest that the model may detect subtle or structural patterns in the ECG predictive of future ischemic events.

Ischemic heart disease prevalence and outcomes are known to be highly variable across patient age, sex, and race/ethnicity^17^. Moreover, studies have demonstrated the need to assess potential bias of analyzing ECGs across diverse demographics^18^. To understand how our model performs across different patient profiles we performed subgroup analyses accordingly. We observed consistent performance across sex and race/ethnicity. Although the model was accurate in patients of all ages, we found performance was best in younger patients. Notably, the model was less accurate in patients with paced rhythms or left bundle branch block (Supplementary Figure 3). However, given the difficulty in identifying subtle ECG abnormalities in patients with paced ECGs and left bundle branch block, despite its reduced accuracy in these patients, the model still provided useful and actionable data.

This study has several limitations. First, this is a retrospective, single-center study that requires further external and prospective validation before translation to the clinical arena. Second, due to sample sizing, the RWMA endpoint defined in our study did not differentiate between the severity of wall motion, and hypokinesia, akinesia, and dyskinesia were treated similarly as “wall motion abnormalities”. Discriminating between patients with hypokinetic, akinetic, and dyskinetic wall motion abnormalities could have additional clinical utility. Third, the echocardiographic findings were dependent on the final read of the interpreting cardiologist, which is known to be subject to interobserver variability. Systematic reinterpretation was not performed due to the impracticality of reinterpreting the large number of studies included.

In conclusion, deep neural networks models can accurately screen for regional wall motion abnormalities and identify patients at increased risk of future ischemic heart disease using a low cost and ubiquitous 12-lead ECG. Based on the observed performance from this study, this model may support and inform clinical decision making for patients with ischemic heart disease who could benefit from medical intervention.

## Data Availability

Data used for the study was collected from three public databases, Medical Information Mart for Intensive Care (MIMIC)-IV-ECG, MIMIC-IV-Note, and MIMIC-IV-ECHO. Clinical data linked to patients included in the study was matched using the publicly available database, MIMIC-IV

https://physionet.org/content/mimic-iv-echo/0.1/

https://physionet.org/content/mimic-iv-ecg/1.0/

---

**Supplementary Figure 1.**
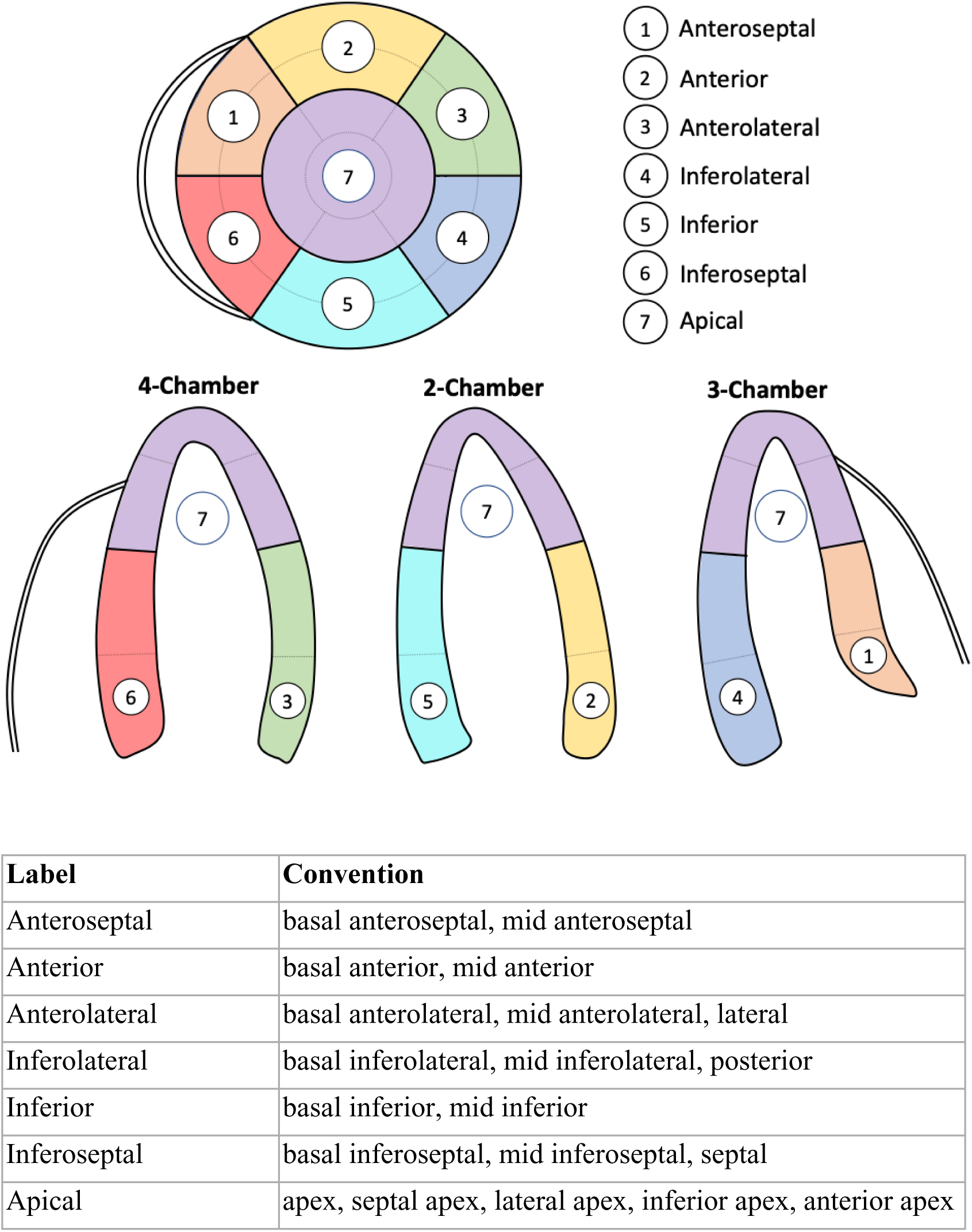
Left ventricular wall segme labeling convention for each of the seven ASE regions^14^.

**Supplementary Table 1.** ECG findings from the entire cohort. Values are provided as n (%). Chi-squared test was used.

|  | RWMA |  |  | RV hypokinesis |  |  | LVEF ≤40% |  |  |
| --- | --- | --- | --- | --- | --- | --- | --- | --- | --- |
|  | Control | Case | P value | Control | Case | P value | Control | Case | P value |
| **Primary rhythm** |  |  |  |  |  |  |  |  |  |
| **Normal sinus rhythm** | 37418 (53.0) | 14810 (58.6) | <0.001 | 43476 (56.4) | 4007 (41.4) | <0.001 | 39684 (55.4) | 11853 (51.7) | <0.001 |
| **Atrial fibrillation** | 11699 (16.6) | 3374 (13.3) | <0.001 | 11107 (14.4) | 2527 (26.1) | <0.001 | 10650 (14.9) | 4176 (18.2) | <0.001 |
| **Sinus tachycardia** | 9828 (13.9) | 2539 (10.0) | <0.001 | 9850 (12.8) | 1205 (12.5) | 0.401 | 9363 (13.1) | 2813 (12.3) | 0.002 |
| **Sinus bradycardia** | 5665 (8.0) | 1906 (7.5) | 0.016 | 6568 (8.5) | 436 (4.5) | <0.001 | 6447 (9.0) | 1054 (4.6) | <0.001 |
| **Atrial flutter** | 1968 (2.8) | 489 (1.9) | <0.001 | 1726 (2.2) | 460 (4.8) | <0.001 | 1714 (2.4) | 701 (3.1) | <0.001 |
| **Atrial tachycardia** | 937 (1.3) | 251 (1.0) | <0.001 | 849 (1.1) | 193 (2.0) | <0.001 | 806 (1.1) | 361 (1.6) | <0.001 |
| **Ectopic atrial rhythm** | 801 (1.1) | 334 (1.3) | 0.02 | 900 (1.2) | 102 (1.1) | 0.357 | 847 (1.2) | 274 (1.2) | 0.906 |
| **Junctional rhythm** | 709 (1.0) | 284 (1.1) | 0.115 | 702 (0.9) | 181 (1.9) | <0.001 | 735 (1.0) | 235 (1.0) | 0.982 |
| **Supraventricular tachycardia** | 572 (0.8) | 160 (0.6) | 0.006 | 509 (0.7) | 127 (1.3) | <0.001 | 516 (0.7) | 208 (0.9) | 0.005 |
| **Ventricular rhythm** | 271 (0.4) | 134 (0.5) | 0.002 | 276 (0.4) | 79 (0.8) | <0.001 | 277 (0.4) | 115 (0.5) | 0.022 |
| **Wide QRS tachycardia** | 160 (0.2) | 120 (0.5) | <0.001 | 169 (0.2) | 70 (0.7) | <0.001 | 101 (0.1) | 173 (0.8) | <0.001 |
| **Ventricular tachycardia** | 126 (0.2) | 112 (0.4) | <0.001 | 151 (0.2) | 54 (0.6) | <0.001 | 79 (0.1) | 150 (0.7) | <0.001 |
| **Junctional ectopic rhythm** | 146 (0.2) | 63 (0.2) | 0.244 | 160 (0.2) | 31 (0.3) | 0.034 | 161 (0.2) | 45 (0.2) | 0.469 |
| **Secondary rhythm** |  |  |  |  |  |  |  |  |  |
| **Premature ventricular complex(es)** | 5634 (8.0) | 2923 (11.6) | <0.001 | 6134 (8.0) | 1544 (16.0) | <0.001 | 4980 (7.0) | 3406 (14.9) | <0.001 |
| **Premature supraventricular complex(es)** | 4356 (6.2) | 1449 (5.7) | 0.013 | 4615 (6.0) | 538 (5.6) | 0.105 | 4361 (6.1) | 1365 (6.0) | 0.466 |
| **Axis deviation** |  |  |  |  |  |  |  |  |  |
| **Left axis deviation** | 6551 (9.3) | 2877 (11.4) | <0.001 | 7291 (9.5) | 1142 (11.8) | <0.001 | 6376 (8.9) | 2933 (12.8) | <0.001 |
| **Right axis deviation** | 1442 (2.0) | 386 (1.5) | <0.001 | 1135 (1.5) | 512 (5.3) | <0.001 | 1369 (1.9) | 431 (1.9) | 0.783 |
| **Atrioventricular conduction delay** |  |  |  |  |  |  |  |  |  |
| **First-degree AV block** | 5568 (7.9) | 2630 (10.4) | <0.001 | 6373 (8.3) | 997 (10.3) | <0.001 | 5653 (7.9) | 2437 (10.6) | <0.001 |
| **High-grade AV block** | 252 (0.4) | 84 (0.3) | 0.615 | 226 (0.3) | 65 (0.7) | <0.001 | 222 (0.3) | 111 (0.5) | <0.001 |
| **Complete heart block** | 135 (0.2) | 48 (0.2) | 0.964 | 141 (0.2) | 24 (0.2) | 0.205 | 140 (0.2) | 42 (0.2) | 0.778 |
| **Ventricular hypertrophy** |  |  |  |  |  |  |  |  |  |
| **Left ventricular hypertrophy** | 6192 (8.8) | 2496 (9.9) | <0.001 | 7206 (9.3) | 818 (8.5) | 0.005 | 6099 (8.5) | 2473 (10.8) | <0.001 |
| **Right ventricular hypertrophy** | 321 (0.5) | 54 (0.2) | <0.001 | 207 (0.3) | 135 (1.4) | <0.001 | 307 (0.4) | 64 (0.3) | 0.002 |
| **Intraventricular conduction delay** |  |  |  |  |  |  |  |  |  |
| **Right bundle branch block** | 5676 (8.0) | 2496 (9.9) | <0.001 | 5905 (7.7) | 1344 (13.9) | <0.001 | 5839 (8.2) | 2191 (9.6) | <0.001 |
| **Left anterior fascicular block** | 4425 (6.3) | 1985 (7.9) | <0.001 | 4898 (6.3) | 815 (8.4) | <0.001 | 4445 (6.2) | 1859 (8.1) | <0.001 |
| **Idioventricular rhythm** | 3569 (5.1) | 2116 (8.4) | <0.001 | 4116 (5.3) | 935 (9.7) | <0.001 | 3095 (4.3) | 2469 (10.8) | <0.001 |
| **Left bundle branch block** | 3316 (4.7) | 2022 (8.0) | <0.001 | 4056 (5.3) | 715 (7.4) | <0.001 | 2332 (3.3) | 2890 (12.6) | <0.001 |
| **Left posterior fascicular block** | 166 (0.2) | 68 (0.3) | 0.386 | 155 (0.2) | 51 (0.5) | <0.001 | 168 (0.2) | 66 (0.3) | 0.181 |
| **Waveform abnormalities** |  |  |  |  |  |  |  |  |  |
| **Non-specific ST-T abnormality** | 17784 (25.2) | 5192 (20.5) | <0.001 | 18602 (24.1) | 2260 (23.4) | 0.109 | 18094 (25.3) | 4607 (20.1) | <0.001 |
| **Artifact** | 7733 (10.9) | 2348 (9.3) | <0.001 | 8175 (10.6) | 951 (9.8) | 0.022 | 7843 (11.0) | 2099 (9.2) | <0.001 |
| **ST-T abnormality** | 7112 (10.1) | 2957 (11.7) | <0.001 | 8252 (10.7) | 881 (9.1) | <0.001 | 7641 (10.7) | 2304 (10.0) | 0.008 |
| **P wave abnormality** | 5934 (8.4) | 2442 (9.7) | <0.001 | 6620 (8.6) | 967 (10.0) | <0.001 | 5798 (8.1) | 2431 (10.6) | <0.001 |
| **QTc prolongation** | 4314 (6.1) | 1838 (7.3) | <0.001 | 4929 (6.4) | 663 (6.9) | 0.082 | 4491 (6.3) | 1587 (6.9) | <0.001 |
| **ST elevation** | 1543 (2.2) | 2062 (8.2) | <0.001 | 2833 (3.7) | 329 (3.4) | 0.191 | 2172 (3.0) | 1363 (5.9) | <0.001 |
| **Myocardial ischemia** |  |  |  |  |  |  |  |  |  |
| **MI of indeterminate age** | 6205 (8.8) | 4897 (19.4) | <0.001 | 8442 (10.9) | 1396 (14.4) | <0.001 | 7009 (9.8) | 3867 (16.9) | <0.001 |
| **Ischemia pattern** | 4679 (6.6) | 3018 (11.9) | <0.001 | 6019 (7.8) | 927 (9.6) | <0.001 | 5132 (7.2) | 2469 (10.8) | <0.001 |
| **Old myocardial infarction** | 4488 (6.4) | 2819 (11.2) | <0.001 | 5588 (7.2) | 949 (9.8) | <0.001 | 4742 (6.6) | 2416 (10.5) | <0.001 |
| **Subacute myocardial infarction** | 3535 (5.0) | 2693 (10.7) | <0.001 | 4797 (6.2) | 775 (8.0) | <0.001 | 3906 (5.5) | 2194 (9.6) | <0.001 |
| **Pacemaker** |  |  |  |  |  |  |  |  |  |
| **Ventricular pacemaker** | 2530 (3.6) | 1521 (6.0) | <0.001 | 2750 (3.6) | 836 (8.6) | <0.001 | 1943 (2.7) | 1985 (8.7) | <0.001 |
| **Atrial pacemaker** | 736 (1.0) | 390 (1.5) | <0.001 | 857 (1.1) | 138 (1.4) | 0.007 | 709 (1.0) | 402 (1.8) | <0.001 |

**Supplementary Table 2.**
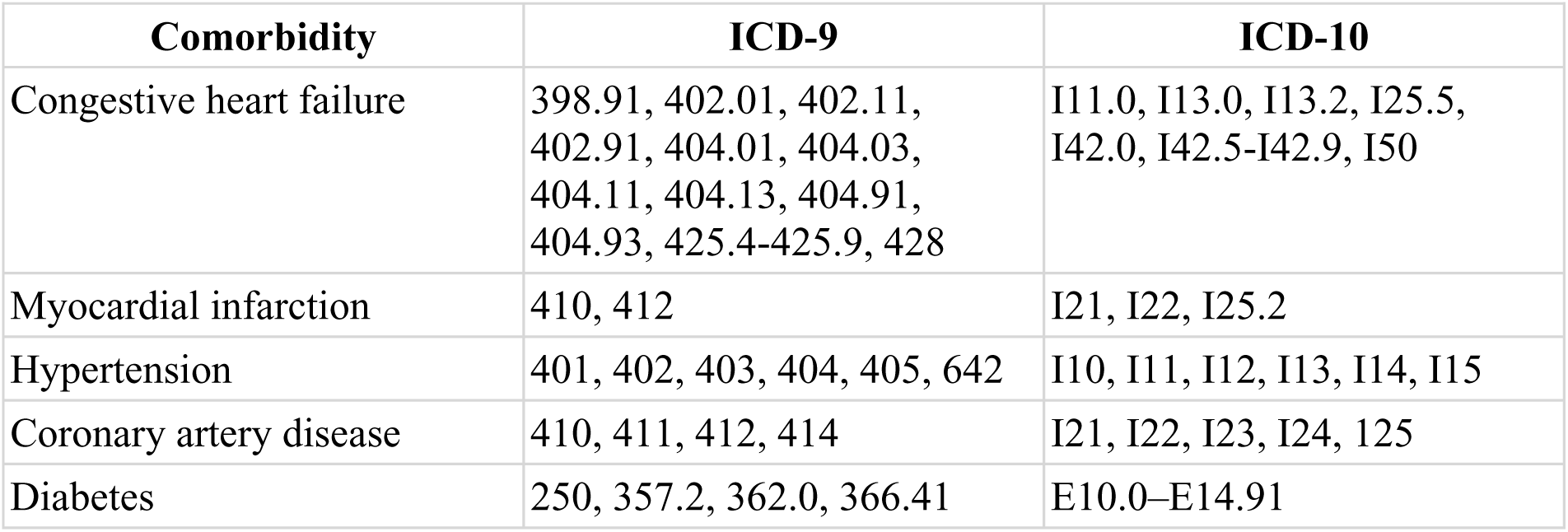
ICD-9 and ICD-10 codes used for MIMIC-IV.

**Supplementary Table 3.** Number of wall motion findings by left ventricular region across the entire cohort.

|  | **Normal** | **Hypokinesis** | **Akinesis** | **Dyskinesis** |
| --- | --- | --- | --- | --- |
| **Anterior wall** | 23192 (93.6%) | 954 (3.8%) | 580 (2.3%) | 64 (0.3%) |
| **Anteroseptal wall** | 22944 (92.6%) | 1214 (4.9%) | 594 (2.4%) | 38 (0.2%) |
| **Inferoseptal wall** | 23114 (93.2%) | 1304 (5.3%) | 355 (1.4%) | 17 (0.1%) |
| **Anterolateral wall** | 23790 (96.0%) | 881 (3.6%) | 118 (0.5%) | 1 (0.0%) |
| **Inferolateral wall** | 22096 (89.1%) | 1622 (6.5%) | 1003 (4.0%) | 69 (0.3%) |
| **Inferior wall** | 21455 (86.5%) | 1886 (7.6%) | 1336 (5.4%) | 113 (0.5%) |
| **Apex** | 21931 (88.5%) | 1528 (6.2%) | 966 (3.9%) | 365 (1.5%) |

**Supplementary Table 4.**
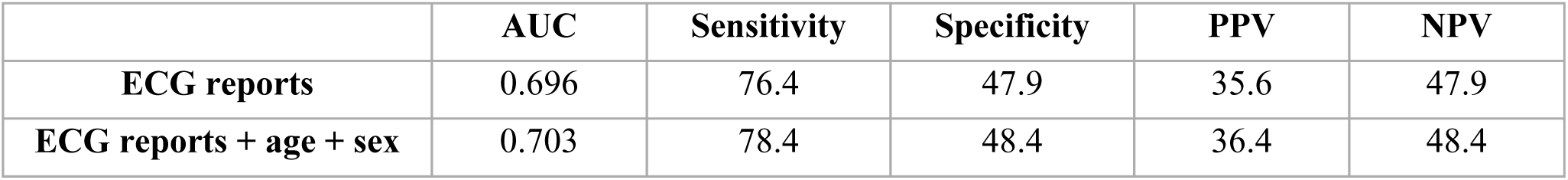
Performance of the reference model on the test set to identify the presence of RWMA.

|  | **AUC** | **Sensitivity** | **Specificity** | **PPV** | **NPV** |
| --- | --- | --- | --- | --- | --- |
| **ECG reports** | 0.696 | 76.4 | 47.9 | 35.6 | 47.9 |
| **ECG reports + age + sex** | 0.703 | 78.4 | 48.4 | 36.4 | 48.4 |

**Supplementary Figure 2.**
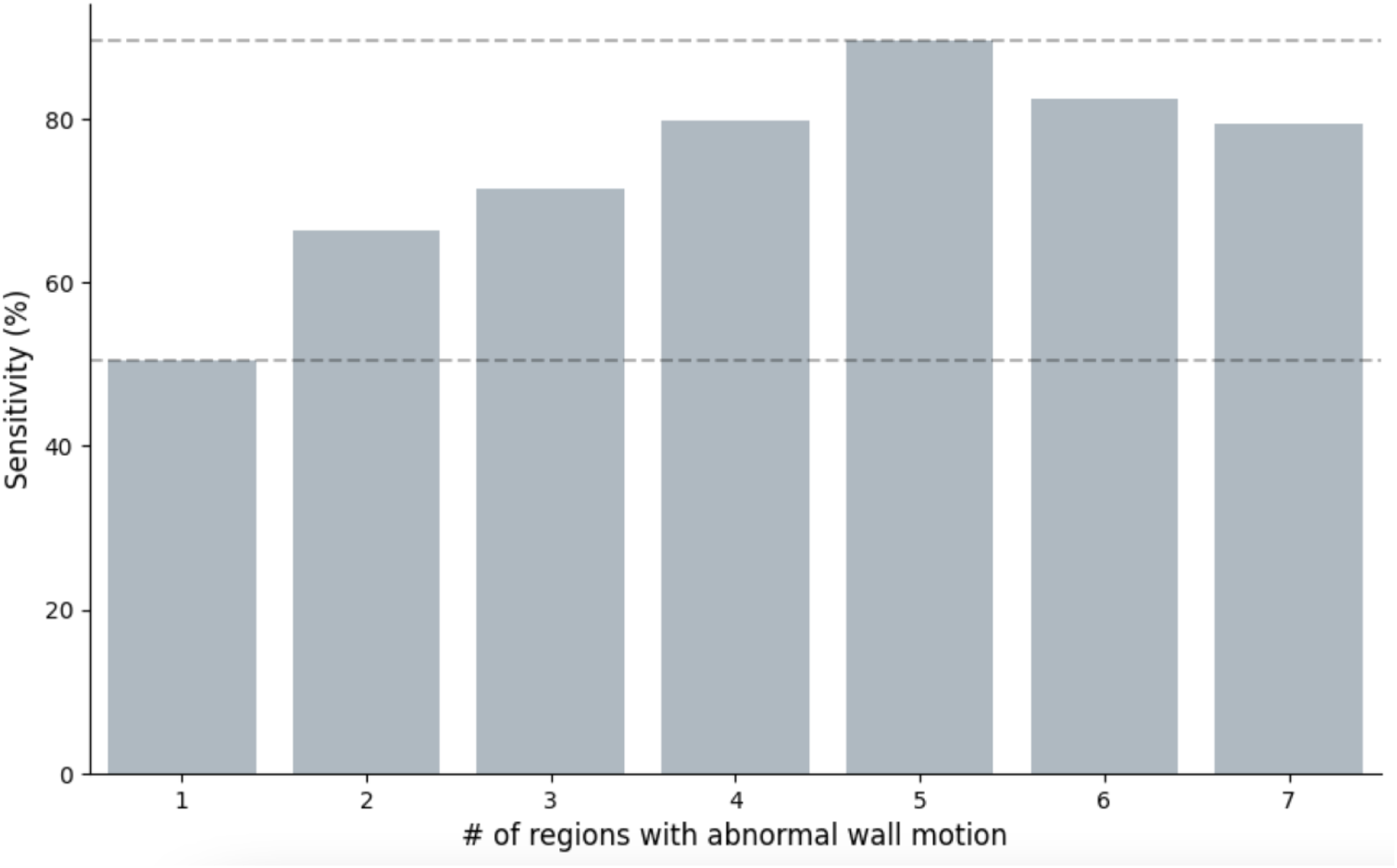
Sensitivity of the deep learning model on the test set to identify the presence of RWMA based on the number of regions with abnormal wall motion.

**Supplementary Figure 3.**
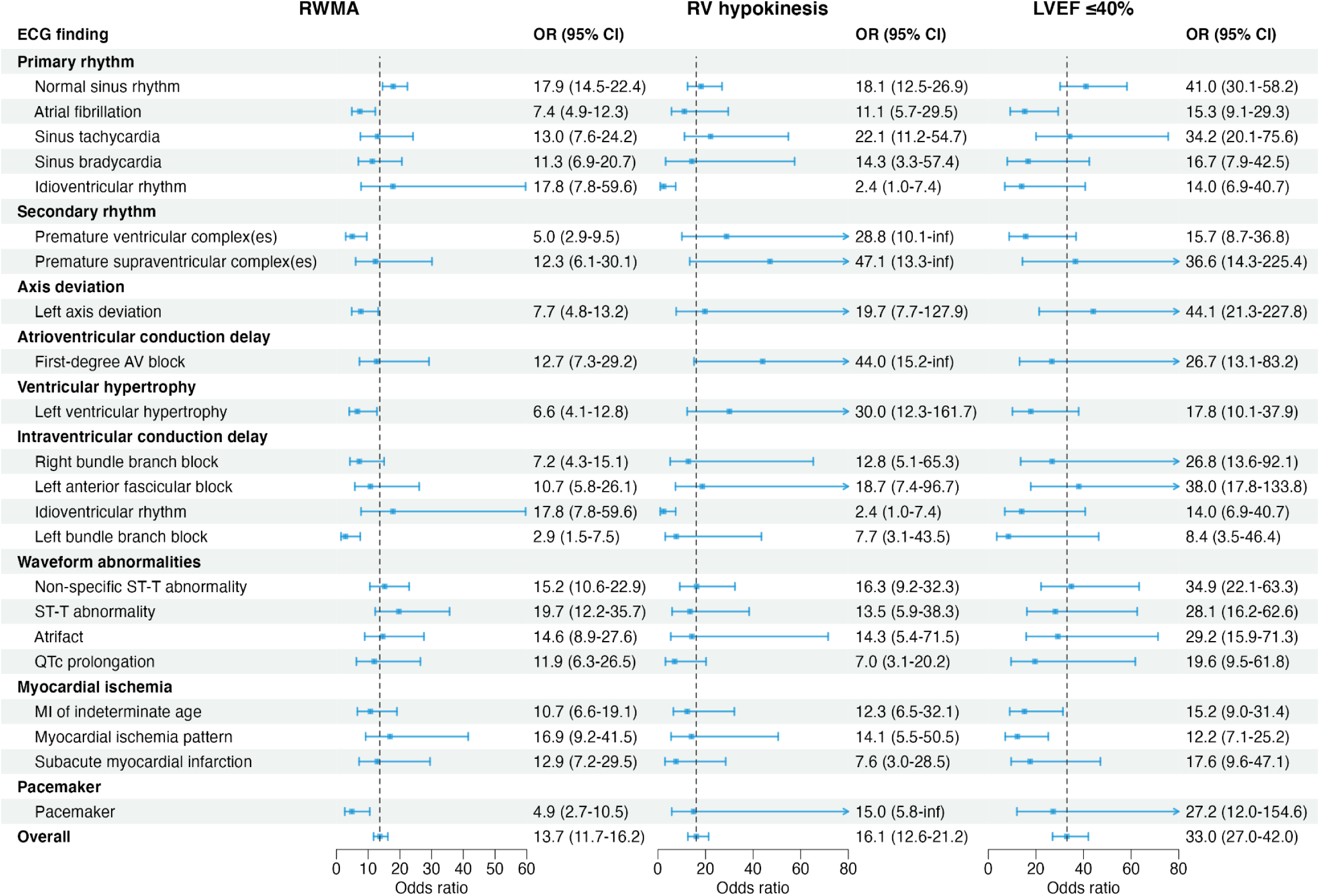
Subgroup analysis by ECG findings extracted from cardiologist reports in MIMIC-IV-ECG using diagnostic odds ratio (OR) with 95% confidence intervals (CIs). The vertical dashed lines represent the OR of the deep learning model across all patients in the test set.

## Appendix

### Training details

The deep learning model was pre-trained using 235,568 ECGs from MIMIC-IV-ECG with weights randomly initialized using truncated normal initialization (Figure 1). We used the Adam optimizer with an initial learning rate of 3×10–4, ℓ2-weight decay of 1×10–4, default coefficients of β1=0.9 and β2=0.999, and a cosine annealing learning rate scheduler. To reduce model overfit we used stochastic weighted averaging [Izmailov et al.] at each residual block and a dropout layer with P=0.5 for the fully connected layers. The model was trained using binary cross-entropy loss for multilabel classification with label smoothing of 0.05 for each output class. To improve generalizability we applied data augmentation by using random shifting, scaling, gaussian noise, CutOut [DeVries et al.], and lead dropout to the waveform. Fivefold cross-validation was used to assess robustness of the model during development. Models were trained for 100 epochs using an early stopping of 12 consecutive epochs with no improvement to the cross-validation macro AUC. The model with the highest validation AUC on the final cross-validation fold was selected for testing.

## References

1. Tsao, C. W. et al. Heart Disease and Stroke Statistics-2023 Update: A Report From the American Heart Association. Circulation 147, e93–e621 (2023).

2. Townsend, N. et al. Epidemiology of cardiovascular disease in Europe. Nat Rev Cardiol 19, 133–143 (2022).

3. Fihn, S. D. et al. 2014 ACC/AHA/AATS/PCNA/SCAI/STS focused update of the guideline for the diagnosis and management of patients with stable ischemic heart disease: a report of the American College of Cardiology/American Heart Association Task Force on Practice Guidelines, and the American Association for Thoracic Surgery, Preventive Cardiovascular Nurses Association, Society for Cardiovascular Angiography and Interventions, and Society of Thoracic Surgeons. J Thorac Cardiovasc Surg 149, e5–23 (2015).

4. Bracey, A. et al. FOCUS may detect wall motion abnormalities in patients with ACS. Am J Emerg Med 69, 17–22 (2023).

5. Attia, Z. I. et al. Screening for cardiac contractile dysfunction using an artificial intelligence-enabled electrocardiogram. Nat Med 25, 70–74 (2019).

6. Vaid, A. et al. Using Deep-Learning Algorithms to Simultaneously Identify Right and Left Ventricular Dysfunction From the Electrocardiogram. JACC Cardiovasc Imaging 15, 395–410 (2022).

7. Smith, S. W. et al. A deep neural network learning algorithm outperforms a conventional algorithm for emergency department electrocardiogram interpretation. Journal of Electrocardiology 52, 88–95 (2019).

8. Elias, P. et al. Deep Learning Electrocardiographic Analysis for Detection of Left-Sided Valvular Heart Disease. J Am Coll Cardiol 80, 613–626 (2022).

9. Gow, B. et al. MIMIC-IV-ECG: Diagnostic Electrocardiogram Matched Subset. PhysioNet 10.13026/4NQG-SB35.

10. Johnson, A., Pollard, T., Horng, S., Celi, L. A. & Mark, R. MIMIC-IV-Note: Deidentified free-text clinical notes. PhysioNet 10.13026/1N74-NE17.

11. Gow, B. et al. MIMIC-IV-ECHO: Echocardiogram Matched Subset. PhysioNet 10.13026/EF48-V217.

12. Johnson, A. E. W. et al. MIMIC-IV, a freely accessible electronic health record dataset. Sci Data 10, 1 (2023).

13. Cerqueira, M. D. et al. Standardized myocardial segmentation and nomenclature for tomographic imaging of the heart. A statement for healthcare professionals from the Cardiac Imaging Committee of the Council on Clinical Cardiology of the American Heart Association. Circulation 105, 539–542 (2002).

14. Lang, R. M. et al. Recommendations for chamber quantification: a report from the American Society of Echocardiography’s Guidelines and Standards Committee and the Chamber Quantification Writing Group, developed in conjunction with the European Association of Echocardiography, a branch of the European Society of Cardiology. J Am Soc Echocardiogr 18, 1440–1463 (2005).

15. Bozkurt, B. et al. Universal definition and classification of heart failure: a report of the Heart Failure Society of America, Heart Failure Association of the European Society of Cardiology, Japanese Heart Failure Society and Writing Committee of the Universal Definition of Heart Failure: Endorsed by the Canadian Heart Failure Society, Heart Failure Association of India, Cardiac Society of Australia and New Zealand, and Chinese Heart Failure Association. Eur J Heart Fail 23, 352–380 (2021).

16. He, K., Zhang, X., Ren, S. & Sun, J. Deep Residual Learning for Image Recognition. in 2016 IEEE Conference on Computer Vision and Pattern Recognition (CVPR) 770–778 (2016). doi:10.1109/CVPR.2016.90.

17. McClellan, M., Brown, N., Califf, R. M. & Warner, J. J. Call to Action: Urgent Challenges in Cardiovascular Disease: A Presidential Advisory From the American Heart Association. Circulation 139, e44–e54 (2019).

18. Macfarlane, P. W., McLaughlin, S. C., Devine, B. & Yang, T. F. Effects of age, sex, and race on ECG interval measurements. J Electrocardiol 27 Suppl, 14–19 (1994).

## Appendix references

Izmailov, P., Podoprikhin, D., Garipov, T., Vetrov, D., & Wilson, A. G. (2018). Averaging weights leads to wider optima and better generalization. arXiv preprint arXiv:1803.05407.

DeVries, T., & Taylor, G. W. (2017). Improved regularization of convolutional neural networks with cutout. arXiv preprint arXiv:1708.04552.

